# “Having a newborn is stressful enough:” Mothers’ experiences during the 2022 Infant Formula Shortage in Washington D.C.

**DOI:** 10.1101/2022.07.31.22278224

**Authors:** Allison C. Sylvetsky, Sarah A. Hughes, Hailey R. Moore, Jeanne Murphy, Janae T. Kuttamperoor, Jennifer Sacheck, Emily R. Smith

**Author notes:** Corresponding author: Allison C. Sylvetsky, PhD, 950 New Hampshire Avenue NW, Suite 200, Washington D.C. 20052.

## Abstract

**Objective:** To investigate mothers’ experiences during the 2022 infant formula shortage in the United States and its perceived impacts on infants’ diet and health.

**Methods:** Mothers of infants under 8 months old were recruited from Washington D.C. using social media and neighborhood listservs and invited to participate in a virtual study meeting between June 22 and July 14, 2022. Mothers completed a brief survey with questions about their demographic characteristics, infants’ anthropometric characteristics, and infant feeding practices, and participated in an in-depth, qualitative interview about their experiences during the infant formula shortage. Survey data were analyzed using means and frequencies, as appropriate. Qualitative interviews were recorded, transcribed verbatim, coded, and thematically analyzed.

**Results:** The sample (n=28) was predominantly White and highly educated. Five overarching themes were identified, including the shortage had: 1) adverse impacts on mothers’ mental and emotional health; 2) significant financial and intangible costs; and 3) led to changes in infant feeding practices; and, 4) social and family networks were helpful in navigating the shortage; and 5) mothers felt fortunate to have resources to breastfeed and/or obtain formula. Most mothers reported the shortage had not yet detrimentally impacted their infant’s health.

**Conclusions:** Even among highly educated women with access to financial, social, structural resources, the infant formula shortage adversely impacted mothers’ mental and emotional health, and has been costly, in terms of financial and intangible costs. Findings demonstrate the urgent need to develop strategies to support mothers in feeding their infants, especially mothers who may lack the resources to locate and obtain formula.

## Introduction

An unprecedented shortage of infant formula occurred in the United States (US) in 2022. This is alarming because the vast majority of US infants are partially or entirely reliant on infant formula for nutrition, with only one in four infants exclusively breastfed for the first six months of life.^1^ The infant formula shortage was caused in part by voluntary recalls of several formulas produced by Abbott Nutrition, shutdown of an Abbott Nutrition plant in Michigan in February 2022 due to bacterial contamination,^2^ and ongoing supply chain interruptions resulting from the COVID-19 pandemic.^3^ Because Abbott is among the largest suppliers of infant formula in the US, and produces specialized formulas for infants with severe allergies, gastrointestinal conditions, and metabolic disorders,^4^ access to specialized formulas rapidly diminished early in 2022. This resulted in a lack of appropriate nutrition for infants with these and other health conditions.^5^ In the months to follow, the availability of more routine formulas also declined drastically, posing widespread challenges to infant feeding nationwide.^5^ The purpose of this qualitative study was to investigate mothers’ experiences during the infant formula shortage and to examine perceived impacts of the shortage on infants’ diet and health. The findings reported herein are from interviews conducted with predominantly non-Hispanic White and highly educated mothers, and will be updated following completion of an ongoing second phase of interviews with a more diverse population, including low-income mothers, in Washington D.C.

## Materials and Methods

In-depth qualitative interviews were conducted virtually (via Zoom™) with mothers of infants under 8 months of age. Mothers from throughout Washington D.C. were recruited using community listservs and social media groups. Interested mothers were contacted by the research team to determine study eligibility. Inclusion criteria included mothers’ report that they: (1) resided in Washington D.C. and (2) had a child less than 8 months old. Recruitment took place from June 20, 2022 to July 11, 2022, and interviews occurred between June 22, 2022 and July 14, 2022. All study procedures were reviewed and approved by the Institutional Review Board at the George Washington University (IRB #: NCR224282).

Eligible mothers were scheduled for a virtual study meeting with a research team member (ACS), which took place in a private Zoom™ room. Mothers provided informed consent via RedCap™, prior to completing a brief survey. The survey included questions about the mother’s and infant’s demographics, infant anthropometrics, how the mother was feeding their infant, and what types of information the mother received about infant feeding. Following completion of the survey, an in-depth interview was conducted by a trained interviewer (ACS) using a semi-structured guide developed collaboratively by the research team. The guide included questions about mothers’ feelings related to the infant formula shortage, how difficult it was to find formula during the shortage, the extent to which mothers perceived the infant formula shortage impacted their infants’ diet, weight, and health, and how the shortage affected their experience as the mother of an infant. All interviews were recorded and transcribed verbatim using NVivo Transcription. Each mother received $50 as compensation for participation.

Descriptive statistics were used to summarize participants’ demographic information and survey responses. Three coders (SAH, HRM, and JTK) independently coded an initial subset of three transcripts using Microsoft Word, which formed the basis for creation of a shared codebook. The remaining transcripts were then coded by two coders using the shared codebook. The coders added new codes as they emerged and reorganized existing codes, as appropriate, to develop the final codebook. All transcripts were subsequently reviewed to ensure that coding was performed in accordance with the final codebook, and any discrepancies were resolved through discussion amongst the coders, with assistance from the interviewer (ACS). Emergent themes and subthemes were then identified, discussed amongst the research team, and refined collaboratively. Representative quotations for each theme and subtheme were selected.

## Results

Twenty-eight mothers enrolled in this mixed-method study. The sample was predominantly White (68%) and highly educated, with 64% of the participants reporting completion of a graduate or doctorate degree (**Table 1**). Two mothers reported their household received nutrition assistance through SNAP (Supplemental Nutrition Assistance Program) and/or WIC (Women, Infants, and Children). The mean age of the infants was 4.6 months. Approximately half reported their infant was female (54% reported having a girl, 46% reported having a boy) and about two-thirds reported that they were using infant formula, whether exclusively (26%) or in combination with breast milk (37%). Of those using formula, over half (59%) reported that they had to switch the brand and/or type of formula they provided to their infant due to the shortage.

**Table 1.**
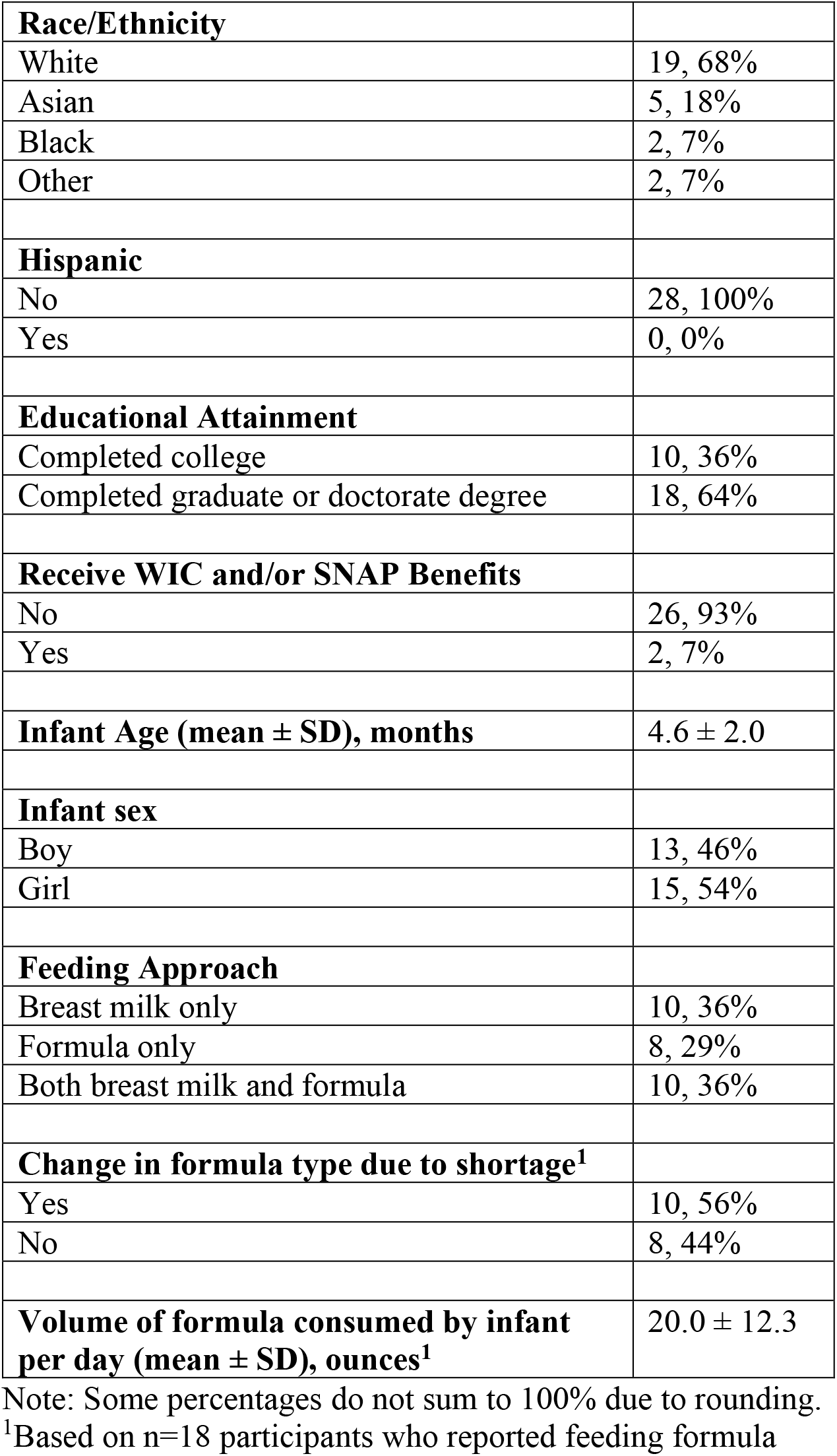
Participant demographics and infant feeding practices, n=28.

A key overarching theme was the infant formula shortage adversely impacted mothers’ mental and emotional health (**Table 2**). Mothers experienced significant anxiety related to the infant formula shortage, specifically surrounding the prospect of not being able to find formula as well as fear that other mothers would be unable to locate formula. They also described feelings of shock and disbelief that a formula shortage could happen in the US. Even mothers who were exclusively breastfeeding reported considerable anxiety about the possibility of having to rely on infant formula, given the possibility that it may not be available. Mothers explained that they had feelings of general stress about the shortage, as well as anger about the situation. Guilt about feeding their infant formula and heightened pressure to breastfeed were also described.

**Table 2.** The infant formula shortage adversely impacted mothers’ mental and emotional health.

| **Theme**<br>**Subtheme** | **Selected Relevant Quotations** |
| --- | --- |
| **Stress** | Having an infant is stressful enough as it is, and having to deal with formula shortages - it's really, really scary.<br>Obviously having an infant is difficult and this has just proven to be one additional stressor on top of a very stressful time in our country. |
| **Anger** | In this country at this time, the fact that we can say it is hard to safely feed your baby when clearly all they want us to do is crank out babies is mind boggling and infuriating, and just makes you want to burn things down.<br>You shouldn't have to worry about whether or not you can feed your baby. It's just horrific. |
| **Shock** | Especially when the news was just coming out; having a newborn, it just felt crazy that we could end up in a situation like that.<br>I mean, we are a developed country. It's mind boggling that this is happening. |
| **Anxiety** | I don't think anybody would have foreseen the sort of emotional distress that comes with worrying that you're not going to be able to feed your baby.<br>It was just the feeling of panic that was [at the end of a day] very predominant. |
| Anxiety about finding formula | It's a big source of stress, and I've tried to keep that anxiety in check. But at times, all of a sudden, it's like, "holy crap, what if I can't feed my baby?" That's scary.<br>My initial thought when I switched to formula was that we wouldn't have any more challenges. I was not prepared to have so much anxiety about it. |
| Anxiety about others having difficulty | I mean, there are definitely families that don't have what they need...it is scary. |
| obtaining formula | A big part of my stress was thinking about all the people who are in different situations and the impact that it would have on them...it's just heartbreaking. |
| Fear about the prospect of having to use formula | It made me scared to think that that I might have to use formula.<br>It has made me scared to attempt to come off breastfeeding earlier and has made me try to really take precautions so that we won't have to rely on formula. |
| Anxiety about milk supply | But when it's [formula] not available, all of a sudden it's like, "OK, I need to get like breastfeeding supplements to try and produce more milk now and make sure that my baby's fed."<br>It has made me more anxious about the supply of breast milk that I have and wanting to make sure that I have enough frozen so that it'll be OK. |
| **Pressure to breastfeed** | I normally hate pumping and it's not something I really want to do. But obviously right now, I just don't really trust the formula situation... I just basically powered through a lot of the nipple pain and stuff like that. I was like, "I can't mess up my breastfeeding right now."<br>Originally, I would have probably stopped pumping by now, but because of the shortage, I have been going a little bit longer. |
| **Guilt about feeding formula** | It made me feel guilty for stopping breastfeeding...I felt a lot of guilt about stopping breastfeeding and all these feelings of being a bad mother for not breastfeeding - they came back.<br>It felt like a failure to have to give some formula in a way that I don't intellectually believe. |

Another key overarching theme was that navigating the infant formula shortage came with significant financial and other intangible costs (**Table 3**). Although most mothers explained that it was not difficult for their family to afford formula, they described spending more money on formula than they had planned, due to the rising cost of formula, the need to purchase formula in larger quantities or “stockpile.” Mothers also described intangible costs, including spending exorbitant amounts of time and energy searching for formula; this entailed driving from store to store, sometimes for hours at a time, and by searching online. Mothers also described sacrificing their physical and emotional health, lifestyle, and sleep to prolong breastfeeding and/or continue pumping to avoid reliance on formula.

**Table 3.**
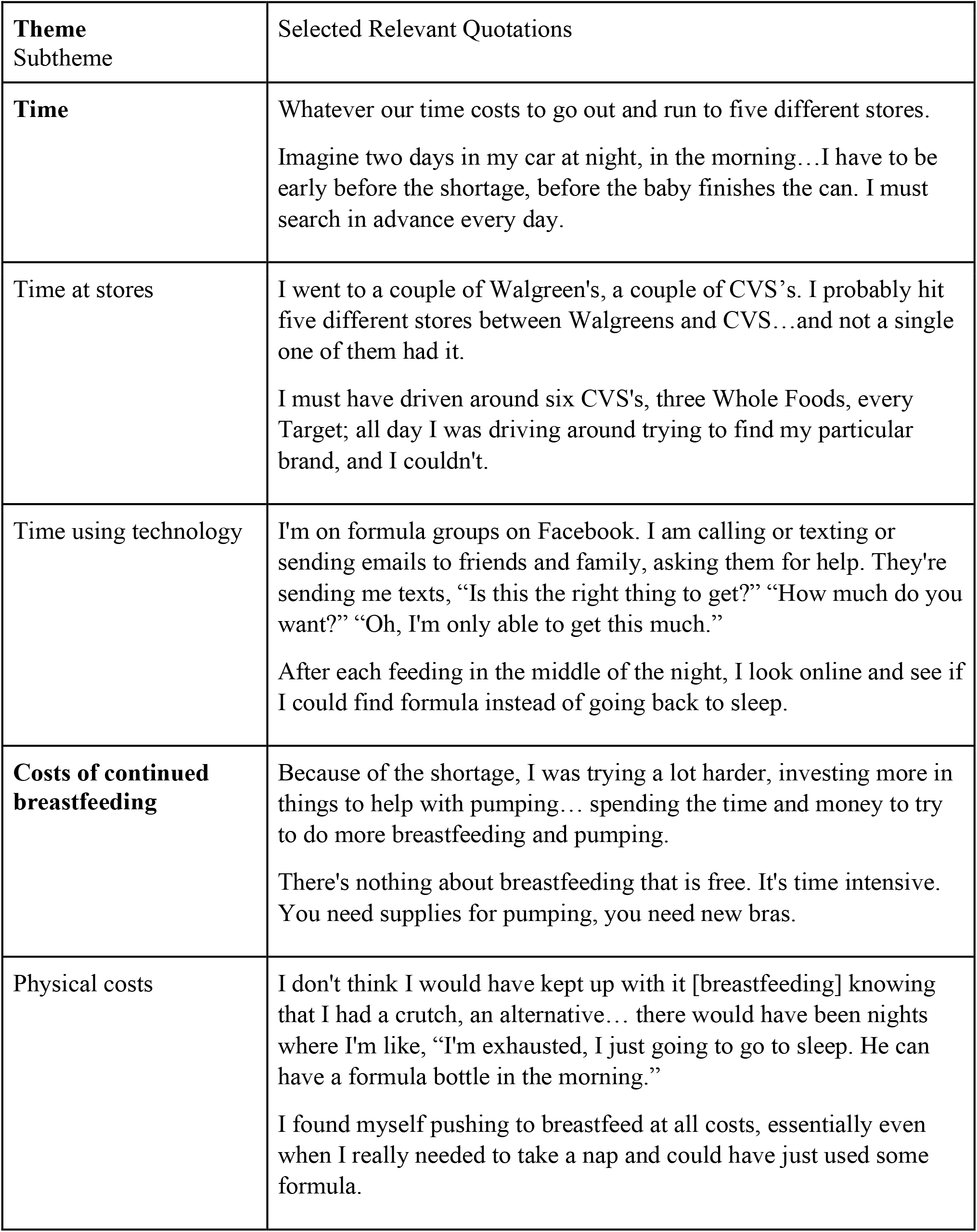
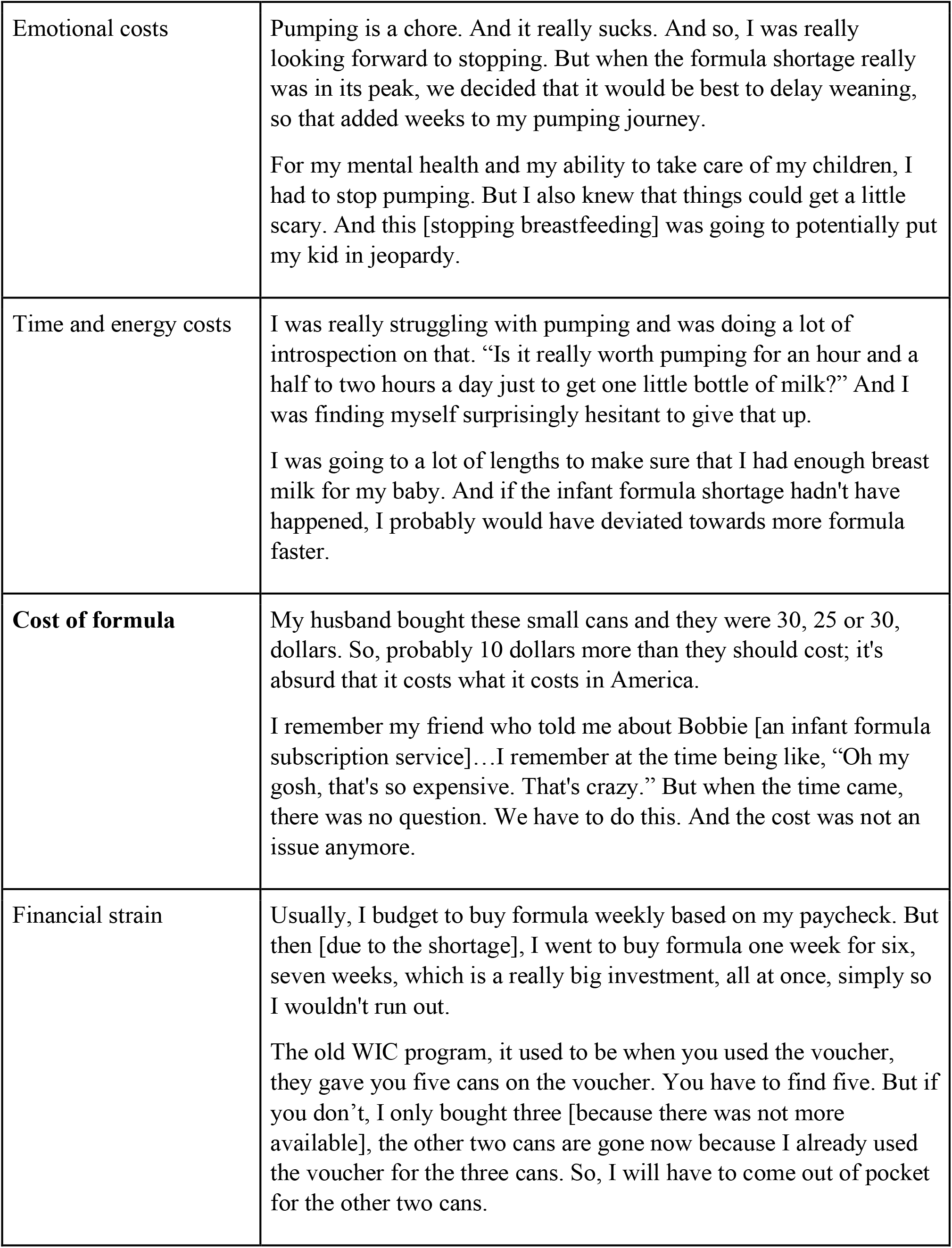
The infant formula shortage had significant financial and intangible costs.

| Theme<br>Subtheme | Selected Relevant Quotations |
| --- | --- |
| **Time** | Whatever our time costs to go out and run to five different stores.<br>Imagine two days in my car at night, in the morning...I have to be early before the shortage, before the baby finishes the can. I must search in advance every day. |
| Time at stores | I went to a couple of Walgreen's, a couple of CVS's. I probably hit five different stores between Walgreens and CVS...and not a single one of them had it.<br>I must have driven around six CVS's, three Whole Foods, every Target; all day I was driving around trying to find my particular brand, and I couldn't. |
| Time using technology | I'm on formula groups on Facebook. I am calling or texting or sending emails to friends and family, asking them for help. They're sending me texts, "Is this the right thing to get?" "How much do you want?" "Oh, I'm only able to get this much."<br>After each feeding in the middle of the night, I look online and see if I could find formula instead of going back to sleep. |
| **Costs of continued breastfeeding** | Because of the shortage, I was trying a lot harder, investing more in things to help with pumping... spending the time and money to try to do more breastfeeding and pumping.<br>There's nothing about breastfeeding that is free. It's time intensive. You need supplies for pumping, you need new bras. |
| Physical costs | I don't think I would have kept up with it [breastfeeding] knowing that I had a crutch, an alternative... there would have been nights where I'm like, "I'm exhausted, I just going to go to sleep. He can have a formula bottle in the morning."<br>I found myself pushing to breastfeed at all costs, essentially even when I really needed to take a nap and could have just used some formula. |
| Emotional costs | Pumping is a chore. And it really sucks. And so, I was really looking forward to stopping. But when the formula shortage really was in its peak, we decided that it would be best to delay weaning, so that added weeks to my pumping journey.<br>For my mental health and my ability to take care of my children, I had to stop pumping. But I also knew that things could get a little scary. And this [stopping breastfeeding] was going to potentially put my kid in jeopardy. |
| Time and energy costs | I was really struggling with pumping and was doing a lot of introspection on that. "Is it really worth pumping for an hour and a half to two hours a day just to get one little bottle of milk?" And I was finding myself surprisingly hesitant to give that up.<br>I was going to a lot of lengths to make sure that I had enough breast milk for my baby. And if the infant formula shortage hadn't have happened, I probably would have deviated towards more formula faster. |
| Cost of formula | My husband bought these small cans and they were 30, 25 or 30, dollars. So, probably 10 dollars more than they should cost; it's absurd that it costs what it costs in America.<br>I remember my friend who told me about Bobbie [an infant formula subscription service]...I remember at the time being like, "Oh my gosh, that's so expensive. That's crazy." But when the time came, there was no question. We have to do this. And the cost was not an issue anymore. |
| Financial strain | Usually, I budget to buy formula weekly based on my paycheck. But then [due to the shortage], I went to buy formula one week for six, seven weeks, which is a really big investment, all at once, simply so I wouldn't run out.<br>The old WIC program, it used to be when you used the voucher, they gave you five cans on the voucher. You have to find five. But if you don't, I only bought three [because there was not more available], the other two cans are gone now because I already used the voucher for the three cans. So, I will have to come out of pocket for the other two cans. |

A third overarching theme was that the shortage led to changes in infant feeding practices (**Table 4**). Mothers described continuing to breastfeed and pump breast milk, despite having planned to stop breastfeeding, to prevent reliance on and/or conserve formula. They also mentioned freezing extra milk to have a back-up option, and in some cases, donating their extra breast milk to help other families. Mothers also explained that they purchased large quantities of formula at one time to ensure they would have enough if they were subsequently unable to locate formula. Mothers also reported making efforts to conserve formula by reducing how much formula they provided to their baby, saving leftover formula, and encouraging their baby to finish bottles, as well as earlier introduction of solid foods.

**Table 4.** The infant formula shortage led to changes in infants’ diet and gastrointestinal health.

| Theme<br>Subtheme | Selected Relevant Quotations |
| --- | --- |
| **Feeding breast milk instead of formula** | If the formula shortage wouldn't have happened. I would have just started giving her formula much sooner.<br>It's like, "let's not become dependent on something that may or may not be there for us," and so I think that that has meant that instead of supplementing at times we just continue to breastfeed. |
| Freezing more milk | We are freezing a lot more milk...and I'm pumping at work so that we can make sure that when I'm not at home, we have milk to rely on and not formula.<br>We thought if we froze it, then just in case we had an issue with finding formula at some point, we would have a little bit in the freezer, just in case. |
| **Stockpiling formula** | I think we got about 10 containers worth. And each container is good for a couple of weeks, so that's almost half a year's worth of supply at the rate he was drinking at the time.<br>I'm acutely aware that you can't run down the store down the street and just pick it up. You have to have a stockpile. |
| **Conserving formula** | [Prior to the shortage], I would make extra milk in case she would want it, and now I'm super regulated; I'm very protective of that scoop, try to get her on a schedule, no wasted formula, like we can't afford to waste because it's so scary.<br>We have to be careful about how much we give her because we can't just keep pouring out four ounces. |
| Minimizing wasted formula | I probably stretched some of the limits: "there's this much left, okay, we're going to stretch it and go a little bit." I've never gone more than, let's say, two days, but I probably stretch some of the recommended limits.<br>Definitely not wanting to waste milk in a bottle; you know, making sure that he's finishing bottles, and it's frustrating if he doesn't. |
| **Donating milk** | I've actually been donating milk and tried to donate a little more with the formula shortage happening. I've tried to do a little extra pumping; just kind of knowing that the shortage is happening and trying in a small way to help out.<br>I'll probably start donating breast milk, which I wouldn't necessarily have even really thought about if there were not this shortage happening. |
| **Switching type of formula** | I'm kind of hesitant to switch back and forth, but I also don't really have a choice I would need to give her something<br>The biggest impact was really switching from ready-to-feed to the powder. |
| Gastrointestinal problems after switching type of formula | He ended up pooping, an incredible amount, nine times a day for several days. So, the impact is our son was fed, but he wasn't fed the formula that he would normally eat.<br>Switching from ready-to-feed to the powder, there was some gastrointestinal stuff, but nothing that caused me concern. |
| **Earlier introduction of solid foods** | I am probably pushing solids a little bit more than I would be normally, just because it would be great to not be so reliant on formula and get off it quicker than I might have otherwise.<br>I think we would have probably waited [to introduce solid food] until he could have handled things a little bit more and start using softer foods instead of going with all the purees and everything like that. |

Another overarching theme was mothers found social networks helpful in locating and obtaining infant formula, but they did not feel supported by the government **(Table 5)**. Mothers explained that they were able to locate formula through social media groups and had friends and family living in other parts of the country send or bring formula to them. They also described a new or amplified sense of community, where mothers looked out for each other and were unified due to the difficulty of the situation. Meanwhile, mothers expressed frustration at the lack of solutions provided by the government and felt that addressing the formula shortage was not prioritized by politicians.

**Table 5.** Social networks were helpful in navigating the infant formula shortage but mothers did not feel supported by the government.

| Theme<br>Subtheme | Selected Relevant Quotations |
| --- | --- |
| **Support from social networks** | I think it's interesting, the mom's looking out for moms, you know, sending messages about what they're seeing at the grocery store, different stores and stuff...you know, that community of support is nice and is impressive to see.<br>As soon as we started to get low on the ready-to-feed, we reached out to my network. I reached out to my friends. I reached out to my co-workers. I reached out to my family and asked them. And that creates a web up and down the East Coast and across the country. |
| Social media groups | I'm seeing a lot of folks coming out to support each other, like in all these parent groups. I just see so many people posting, "I have extra, do you need this?" Everyone kind of comes together to support each other because of this.<br>You keep seeing everyone posts like, "Oh, I have a couple of extra of this and couple extra of that"...it's great that it's going to good use. |
| Family | My mother and sister are in South Carolina. My mother's retired and my sister doesn't have a traditional office job...once they learned about it [the shortage], they went on a scavenger hunt and were able to get it [our supply of formula] back up.<br>I asked my sister in Colorado to look for the formula as well and say, "Buy what you can. I'll be there in a couple of weeks and then I'll take it back." |
| **Lack of government support** | It started to make the national news after it was already a big problem. And...it's still a big problem. And, the news cycle has moved on. So, I think it's just left parents to continue to try and figure it out themselves.<br>It's kind of like we don't have a government, since the system is set up to step up in this type of moment. |
| Lack of policy solutions | It's frustrating that we're not seeing policy solutions.<br>The politics and laws are not super supportive of moms. |
| Failure to prioritize needs of mothers and infants | The needs of infants and their caregivers are shunted to one side, except when it's a convenient political football.<br>We should be able to put our resources behind it to be able to take care of babies. That should be a priority. But it's not a priority that's getting fixed. |

A final emergent theme was that mothers felt lucky to be able to breastfeed and have resources to obtain formula (**Table 6**). Mothers explained feeling fortunate that their baby was able to latch and/or that their milk supply was plentiful. Mothers also reported feeling grateful that they had resources to obtain formula, including supportive family members and friends, paid maternity leave, financial means to purchase more expensive formula or enroll in formula subscription services, and to live near a plethora of retail outlets that sold infant formula.

**Table 6.** Mothers felt lucky to have the ability to breastfeed and/or obtain formula during the shortage.

|  |  |
| --- | --- |
| **Ability to breastfeed** | I feel really grateful that I am able to feed my baby primarily breast milk, and that I don't have to rely on it [formula], but it is so scary.<br>My body and my baby are cooperating, and it would be a different situation if they weren't. |
| **Ability to obtain formula** | I feel pretty lucky to be of a certain socioeconomic status where we only had trouble for two days.<br>I know that I have relative affluence and I have a big network, so I thought I would be OK because generally, that's the truth. |
| Social resources | I feel lucky that I have the privilege to be able to source formula if I end up needing it from my neighbors.<br>And people who don't have that luxury, don't have a network of people to tap into as their backup plan - it guts me as a mom and as a person. We should all be so angry at this whole thing. |
| Structural resources | We're fortunate...my husband's been on paternity leave, so there's been a couple of mornings where he was out and went to five or six different stores to try and find some.<br>I'm so fortunate because I was in these mom groups. I had maternity leave to go and participate and build this network. |
| Financial resources | It's crazy how much it [formula] costs. We're doing relatively well for ourselves, and I can't imagine what other people are doing. I'm sorry. I think I the last Amazon order I did, it was almost two hundred dollars.<br>We had to get a different brand of formula that was approximately twice as expensive. Because we are fortunate enough to be able to afford it. |
| Access to subscription services | We signed up for a subscription service, so we feel a bit more secure with where we're getting our formula from.<br>We started ordering Bobbie [subscription] formula; and around the same time, like March or April, they put a pause on accepting new customers so that they could preserve their inventory for existing customers. And once I saw that, it took me like a couple of weeks to |
|  | be like, “OK, I feel confident we're still getting the formula we need.” |
| Built environment | I feel grateful that I was able to get like a six, seven-week supply because I scavenger hunt the city.<br>I'm so sorry for the ladies who live in the countryside...they are worse off than us. |

## Discussion

The infant formula shortage adversely impacted mothers’ mental and emotional health; and, navigating the situation had significant and unanticipated financial, physical, emotional, and lifestyle-related (e.g., time) costs. While mothers did not report serious detrimental effects of the shortage on their infants’ health, they commonly explained that they altered their planned and/or preferred feeding practices to ensure their infant received adequate nutrition. However, mothers in the present study were predominantly White and highly educated, and it is likely that the shortage has more significantly impacted women with lower educational attainment and limited financial and/or social resources. Infants from low-income households are also more likely to be reliant on infant formula; 56% of all US infant formula sold each year is consumed by infants enrolled in the Special Supplemental Nutrition Program for Women, Infants, and Children (WIC) program.^6^ This is related to persistent racial/ethnic and socioeconomic disparities in breastfeeding in the US,^7,8^ which result in large part from social determinants including disparities in access to paid maternity leave, lactation support, and breastfeeding education.^9^ In fact, a mother in our sample who was enrolled in WIC described significant financial strain from “wasting” WIC benefits. One WIC voucher purchases five cans of formula, but if fewer than five cans were available, she was forced to use an entire voucher to obtain whatever formula she could. Because this mother could not maximize her WIC benefits, she had to pay for additional formula out of pocket. While this was mentioned only by one participant in this initial sample, it may re-emerge in our ongoing second phase of interviews with mothers from lower-income households.

Adverse consequences of the infant formula shortage on mothers’ mental and emotional health are particularly alarming because the postpartum period is already a time of heightened stress.^10^ In the year following the birth of a child, fifteen percent of women suffer from postpartum anxiety^11^ and approximately 12.5% of women in the US have postpartum depression.^12^ Both conditions have unfavorable short and long-term impacts on maternal health and infant growth^13^ and development,^14^ and also interfere with mother-child bonding^15^ and infant feeding outcomes.^16^ Even in the absence of postpartum anxiety and/or depression, the months following childbirth involve sleep exhaustion, feelings of isolation, new demands of parenting, and changes in physique and sexuality, all of which detract from mothers’ mental and physical health.^17^ The COVID-19 pandemic has further exacerbated mental health challenges during the postpartum period and posed a barrier to postpartum women in accessing medical care and social support.^18^ This is not surprising, as the prevalence of psychiatric illnesses is reported to increase disproportionately among postpartum women during disasters (e.g., earthquakes) and events (e.g., terrorist attacks) relative to other segments of the population.^19^

Difficulty finding formula and the prospect of hypothetically being unable to feed their infant were key contributors to reported increases in stress and anxiety associated with the infant formula shortage. Mothers also described experiencing amplified pressure to breastfeed and heightened feelings of guilt associated with providing infant formula. Even in the absence of an infant formula shortage and global pandemic, feelings of maternal shame and anxiety surrounding infant feeding decisions,^20^ and an appalling lack of social and institutional support for breastfeeding in the US^21^ are well-documented. And even among mothers in our study who reported exclusively breastfeeding, considerable anxiety about their milk supply and ability to avoid switching to formula, due to the shortage, were described. Stress associated with the perception of inadequate milk supply is widespread, and often leads to supplementation with infant formula.^22^ It is particularly notable that this high level of stress surrounding continuation of breastfeeding was described among mothers fortunate to have access to home lactation consultations, lengthy paid maternity leaves, and remote work arrangements.

Mothers whose infants were fed formula described spending exorbitant amounts of time searching for formula, which further aggravated their stress and anxiety associated with the shortage. Time spent searching for formula was reported to disrupt work productivity and detract from time spent with older children. While mothers in the present study were able to devote the necessary time, energy, and resources to locating formula, this may not be feasible for women of lower socioeconomic status; for example, due to a lack of childcare, less flexible work schedules, and/or limited options for transportation. Considering that time constraints are a key barrier to healthy eating and physical activity among postpartum women,^23^ which are important for preventing postpartum weight retention^24^ and psychological morbidity^25,26^, the excessive time spent searching for formula may have indirect, unfavorable impacts on mothers’ physical and psychological health.

Mothers fortunately did not report any serious detrimental effects of the shortage on their infants’ health, apart from adverse gastrointestinal consequences (e.g., diarrhea, gas, discomfort) from switching formula types. The avoidance of more serious infant health problems was predominantly attributed to being able to breastfeed and/or to ultimately finding sufficient formula after taking extreme or unconventional measures to acquire sufficient formula (e.g., leveraging social and family networks, searching store after store to find formula). While reported changes such as conserving formula through saving leftover formula for subsequent feeds and encouraging their infant to finish bottles did not result in major health problems, these practices are inconsistent with federal bottle-feeding guidance^27^ and may pose risks to infants both in terms of food safety,^27^ and contribute to overfeeding.^28^ Furthermore, mothers also reported providing their infant with solid foods earlier than planned, which is concerning because early introduction of solids is associated with later childhood obesity, particularly among formula-fed infants.^29^

Social, partner, and family support were widely described as critical facilitators of acquiring infant formula during the shortage, and are known to positively influence mothers’ physical and mental health during the postpartum period.^30-32^ Mothers explained that they used social media to locate formula and described social media groups as a key source of support in navigating the shortage. Social media is an important source of social support during pregnancy and postpartum,^33,34^ and further efforts to provide postpartum women with support and reputable resources through both online and in-person social networks are warranted. Mothers in our study did not take these networks for granted; however, they explained that the formula shortage made them feel even more fortunate to have the resources and support systems to obtain formula.

A key limitation of this study is enrollment of a predominantly non-Hispanic White and highly educated sample of mothers, who had the financial, social, and educational resources to overcome significant challenges resulting from the infant formula shortage. Findings therefore may not be generalizable to low-income and minority households, or the broader population of postpartum women in the US. It is essential to understand the perspectives of mothers from low-income and/or minority backgrounds, who may have had different experiences during the infant formula shortage. For those living in low-income households, financial and intangible burdens associated with the formula shortage may detrimentally influence household food security and pose a barrier to affording other key necessities such as housing. Although we initially circulated advertisements for this study across Washington D.C., we are now using more targeted recruitment efforts and have narrowed the study eligibility criteria to specifically engage mothers from more diverse and disadvantaged backgrounds in the ongoing, second phase of interviews.

To our knowledge, this is the first study to examine mothers’ experiences during the 2022 infant formula shortage. Findings call attention to concerning impacts of the infant formula shortage on mothers’ mental and emotional health, and underscore the importance of social and family support in locating and acquiring infant formula, as well as the urgent need for policy solutions to address this unprecedented crisis. These initial findings warrant further research to elucidate perceived impacts of the formula shortage among mothers who may lack financial and/or social resources to locate and obtain formula, and will inform development of intervention strategies to ensure that mothers can adequately nourish their infants during this and other unanticipated situations.

## Data Availability

All data produced in the present study are available upon reasonable request to the authors

